# Telemedicine Ready or Not? a cross-sectional assessment of telemedicine maturity of federally funded tertiary health institutions in Nigeria

**DOI:** 10.1101/2022.06.13.22276341

**Authors:** Tolulope F. Olufunlayo, Oluwadamilola O. Ojo, Obianuju B. Ozoh, Osigwe P. Agabi, Chuks R. Opara, Funlola T. Taiwo, Olufemi A. Fasanmade, Njideka U. Okubadejo

**Author notes:** CORRESPONDENCE Dr. Oluwadamilola O. Ojo, Department of Medicine, Faculty of Clinical Sciences, College of Medicine, University of Lagos, and Lagos University Teaching Hospital, Idi-araba, Nigeria.

## Abstract

**Background and Objective:** Telemedicine (TM) has solidified its place as a means for continuity of healthcare services and a cost-effective approach for improving health equity as demonstrated during the COVID-19 pandemic. Preparedness of health systems for telemedicine is an indicator of the scalability of their services, especially during disasters. We aimed to assess the maturity and preparedness of federally funded tertiary health institutions (FFTHIs) in Nigeria, to deploy and integrate telemedicine

**Methods:** We conducted a cross-sectional study of randomly selected FFTHIs in Nigeria using PAHO’s tool for assessing the maturity level of health institutions to implement telemedicine services. Descriptive statistics were used for overall maturity levels and non-parametric tests to compare scores for overall maturity and specific PAHO domains per region. The level of significance was set at p value <0.05.

**Results:** Twenty-four of thirty randomly polled FFTHIs responded (response rate of 77.4%). Overall, the median TM maturity level was 2.0 (1.75) indicating beginner level. No significant inter-zonal difference in median overall maturity level (p=0.87). The median maturity levels for telemedicine readiness in specific domains were organizational readiness - 2.0 (2.0), processes 1.0 (1.0), digital environment 2.0 (3.0), human resources 2.0 (1.0), regulatory issues- 1.5 (1.0) and expertise 2.0 (2.0); mostly at beginner level, with no inter-zonal differences. Most participating institutions had no initiatives in place for domains of processes and regulatory issues.

**Conclusions:** The current status of telemedicine maturity of FFTHIs in Nigeria behoves policy makers to advance the implementation of telemedicine across the country as part of national digital quality healthcare.

## INTRODUCTION

Health care providers and health systems have been inundated by enquiries about strategies to ensure continuity of access to preventive, diagnostic and therapeutic services in the wake of disruptions during the Coronavirus disease 2019 (COVID-19) pandemic. Undoubtedly, the allure of delivery of health care services where patients and providers are separated by distance (telehealth as defined by the World Health Organization) using information and communication technologies has escalated.^1^ The benefits of telehealth beyond bridging the physical divide and enabling improved access include cost effectiveness, improved emergency preparedness, and a decrease in supply-demand mismatch, considering the low ratio of health workforce per population.^2^ At the national level, health service prioritization is driven by extant policies. According to the WHO Global Observatory for eHealth (2015 latest data) in which 123 member states provided responses, 22.0% stated that the country had a specific national telehealth policy, 50.7% had no policy, and 35.0% had no policy but had a reference to telehealth within their national eHealth policy.^3^ The survey identified specific barriers including lack of funding to develop and support telehealth programmes, lack of infrastructure (equipment and/or connectivity), competing health system priorities and a lack of legislation or regulations covering telehealth programs.^3^ The feasibility of leveraging on telehealth services to effectively cushion the negative impact of the disruptions to in-person access to diagnostic and therapeutic services is dependent on the level of maturity of such services. Beyond the pandemic, the advantages of telehealth as a viable alternative platform have made it an increasingly obvious imperative to incorporate telemedicine in sustainable development of health systems. In order to successfully implement telemedicine services, a holistic approach that gives due consideration must be given to the interwoven components such as technology requirements, organizational structures, change management, economic feasibility, societal impacts, perceptions, user-friendliness, evaluation and evidence, legislation, policy and governance.^4^ The primary objective of this exploratory survey was to assess the maturity and level of preparedness of federally funded tertiary health institutions (FFTHIs) in Nigeria to deploy and integrate telemedicine services in providing care to the population. Such data are currently lacking, and are useful as a bench-mark to guide policies aimed at driving improvements in healthcare services or developing services where non-existent.

## METHODS

### Study design and inclusion criteria

We conducted a cross-sectional descriptive study of randomly selected federally funded tertiary institutions across the six geopolitical zones of Nigeria using an electronic survey instrument between 17/9/2020 and 1/9/2021. A database of all federally funded tertiary hospitals (defined as institutions so designated by the Federal Ministry of Health on account of level of care available at the facility) was built from multiple sources including a primary search of the Nigeria Health Facility Registry ^5^ using the filters for facility level (tertiary) and ownership (public). This was further refined using the pre-set inclusion criteria to identify institutions funded by the Federal Government of Nigeria. Ab initio, the plan was to survey at least 50% of the institutions so identified ensuring that a minimum of 3 responses were received from each geopolitical zone. The initial e-mail requesting participation was sent to the Chief Medical Director/Medical Director of the institution with a preceding telephone call to verify contact details. The survey was completed and corroborated by a designated hospital official such as the Chairman Medical Advisory Committee or similarly authorized official with sufficient information to credibly complete the survey. An initial period of 4 – 8 weeks was allowed for responses with e-mail and telephone reminders over an additional 4 – 8-week period on account of anticipated delays occasioned by the ongoing pandemic. One institution whose official refused to participate was replaced.

### Survey instrument and survey processes

The study utilized an electronic survey designed using Google® forms and included a section on basic demographics of the participating institution, and the Pan American Health Organization (PAHO) tool for assessing the maturity level of health institutions to implement telemedicine services (developed by PAHO in collaboration with the World Health Organization (WHO) and the Inter-America Development Bank.^6^ In summary, the instrument is a self-assessment tool that assesses level of maturity to offer telemedicine services, and is scored as none (no initiative in place – 1), beginner (some steps taken, but far from able to implement services – 2), advancing (good progress, and some telemedicine services could begin to be implemented – 3), and ready (everything is ready for telemedicine services to operate at full capacity – 4). In addition, an unscored response option to request expert technical support to make further improvements is included. The component categories of questions within the tool explore six areas of maturity/preparedness: organizational readiness, processes, digital environment, human resources, regulatory issues, and expertise. In addition to the PAHO-specific questions, the preceding section of the survey required the baseline demographics of the institution including the following: location (state, city), hospital bed capacity, patient load (annual in-patient and outpatient load separately) in the preceding three-year period 2017 to 2019, avoiding 2020 due to anticipated issues with data verification and service utilization related to the pandemic restrictions), availability and use of electronic health records system in the institution, demographics of hospital attendees i.e. approximate proportion of paediatrics and adult patient population, clinical departments in the institution, and presence of residency training programs.

### Data analysis

The survey responses were downloaded from the Google ® form as a Microsoft Excel document and analysed using the Statistical Package for Social Sciences (SPSS) version 21 (IBM Corp. Armonk, NY). Descriptive statistics of the baseline characteristics of the participating institutions and the survey responses (maturity level overall and by each category explored i.e., organizational readiness, processes, digital environment, human resources, regulatory issues, and expertise) are presented with appropriate measures of central tendency and dispersion. Non-parametric Kruskal-Wallis test was used to compare median scores for overall maturity (combining all 5 categories) per region with level of significance set at p value <0.05. Available case analysis used in generating overall frequencies of the different maturity scores for the items in the tool.

## RESULTS

### Characteristics of participating institutions

A total of 24 tertiary hospitals of the 30 randomly polled completed the survey, giving a response rate of 77.4%. The institutions’ characteristics are shown in Table 1 and Figure I (geopolitical regional distribution of participating institutions). There were participants from 5 of the 6 geopolitical zones with ∼95% of respondents being residency-training institutions and ∼54% having an in-patient bed capacity between 500 – 1000. The highest proportion of participating institutions came from the North-central geopolitical zone (7, 29.2%), whilst the South-south had the lowest proportion with only two institutions (8.3%). Slightly more than half of participating institutions (54.2%) had a bed capacity of between 501 and 1000; in the preceding three years, median annual in- patient and clinic attendance rates were 10,761.5, and 114,170.5, respectively. The proportion of paediatric to adult hospital attendees was approximately 1:4, and about one-fifth (18%) of all patients seen were above 60 years of age.

**Table 1.** Baseline characteristics of the participating tertiary hospitals

| Characteristic | Summary data |
| --- | --- |
| Institutions offering residency training (n, %) | 23 (95.8%) |
| Annual in-patient attendance in preceding 3 years (median (IQR)) | 10,761.5 (18117.8) |
| Annual clinic attendance in preceding 3 years (median (IQR)) | 114,470.5 (68,720.8) |
| Proportion of hospital patients classified as adults (median (IQR)) | 79.0 (17.8) |
| Proportion of hospital attendees classified as paediatrics (median (IQR)) | 20.0 (19.5) |
| Institutional cut off for defining paediatric age group (median (IQR)) | 16.0 (2.5) |
| Proportion of hospital patients aged >60 years (median (IQR)) | 18.0 (20.3) |
| Institutions using electronic medical records for consultation (n, %) | 9 (37.5%) |
| Institutions with specific budget line for telemedicine (n, %) | 4 (16.7%) |
| Hospital bed capacity |  |
| <500 beds | 11 (45.8%) |
| ≥500 – 1000 beds | 13 (54.2%) |

A little over a third (37.5%) of all institutions used electronic health records for consultation; less than a fifth (16.7%) had a specific budget line for telemedicine. All but one participating institution offered residency training programmes (Table 1).

**Figure I.**
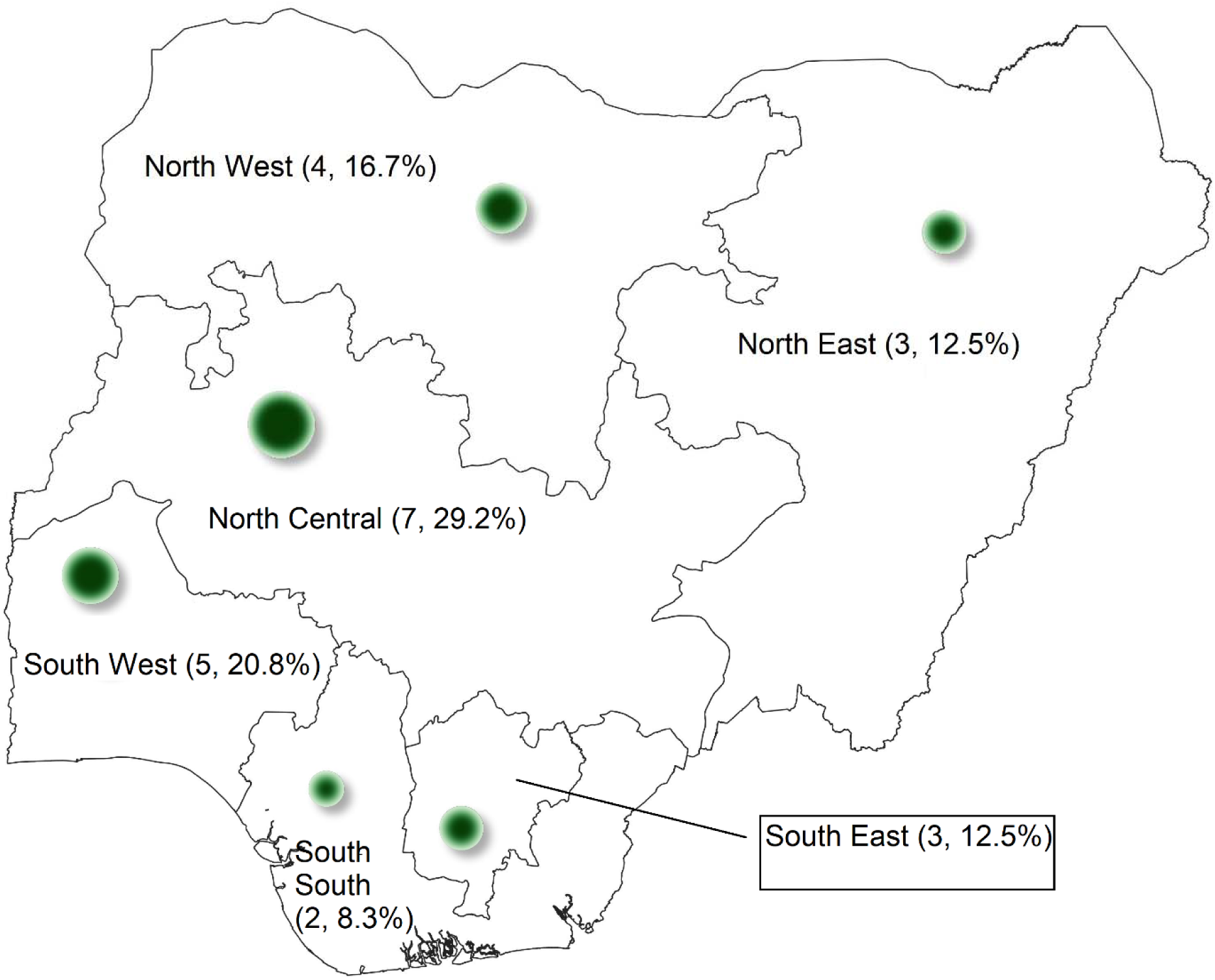
Geopolitical and regional distribution of participating tertiary institutions. Footnote: The green bubbles representing the number and proportion (n, %) of participating institutions in each region. Highest maturity levels per region: NC = 2.0, SE = 2.0, and SW = 2.0. SS = 1.75, NW = 1.5, NE = 1.0). No statistically significant inter- regional difference in median maturity overall (Kruskal Wallis test, p=0.87)

### Telemedicine maturity of tertiary institutions

The summary data for the major thematic areas of the PAHO tool for assessing the maturity level of health institutions to implement telemedicine (Organizational readiness, Processes, Digital environment, Human resources, Regulatory issues, and Expertise) are presented in Table 2 and Figure I (regional data). Supplemental tables 1 – 6 displays the maturity level for all domains and each parameter and the distribution of the different scores among the institutions.

**Table 2.** Institutional maturity for delivery of telemedicine services

| Category | Maturity level | Maturity level |
| --- | --- | --- |
|  | (Median (IQR)) | (Range) |
|  | n = 24 | n=24 |
| Overall maturity level | 2.0 (1.75) | 1 – 4 |
| Organizational readiness | 2.0 (2.0) | 1 – 4 |
| Processes | 1.0 (1.0) | 1 – 4 |
| Digital environment | 2.0 (1.3) | 1 – 4 |
| Human resources | 2.0 (1.0) | 1 – 4 |
| Regulatory issues | 1.5 (1.0) | 1 – 4 |
| Expertise | 2.0 (2.0) | 1 – 4 |

Overall, the median maturity level was 2.0 (1.75) indicating beginner level. There was no statistically significant inter-regional difference in median maturity overall (Figure I, Kruskal Wallis test, p=0.87). Additional details are provided regarding the digital environment (internet connection and connectivity, software for managing medical records and patient portals, administrative software for billing, payments, monitoring hours, technical equipment such as hardware and other equipment) in Table 3 and supplemental table 3. Maturity and level of preparedness in terms of human resource capacity (health workers and IT staff) is depicted in Figure II and supplemental table 4. There was no regional difference in the maturity level for any of the domains (non-parametric test; organizational readiness p=0.37, processes p=0.52, digital environment p=0.86, human resources p=0.88, regulatory issues p=0.28, expertise p=0.10).

**Figure II.**
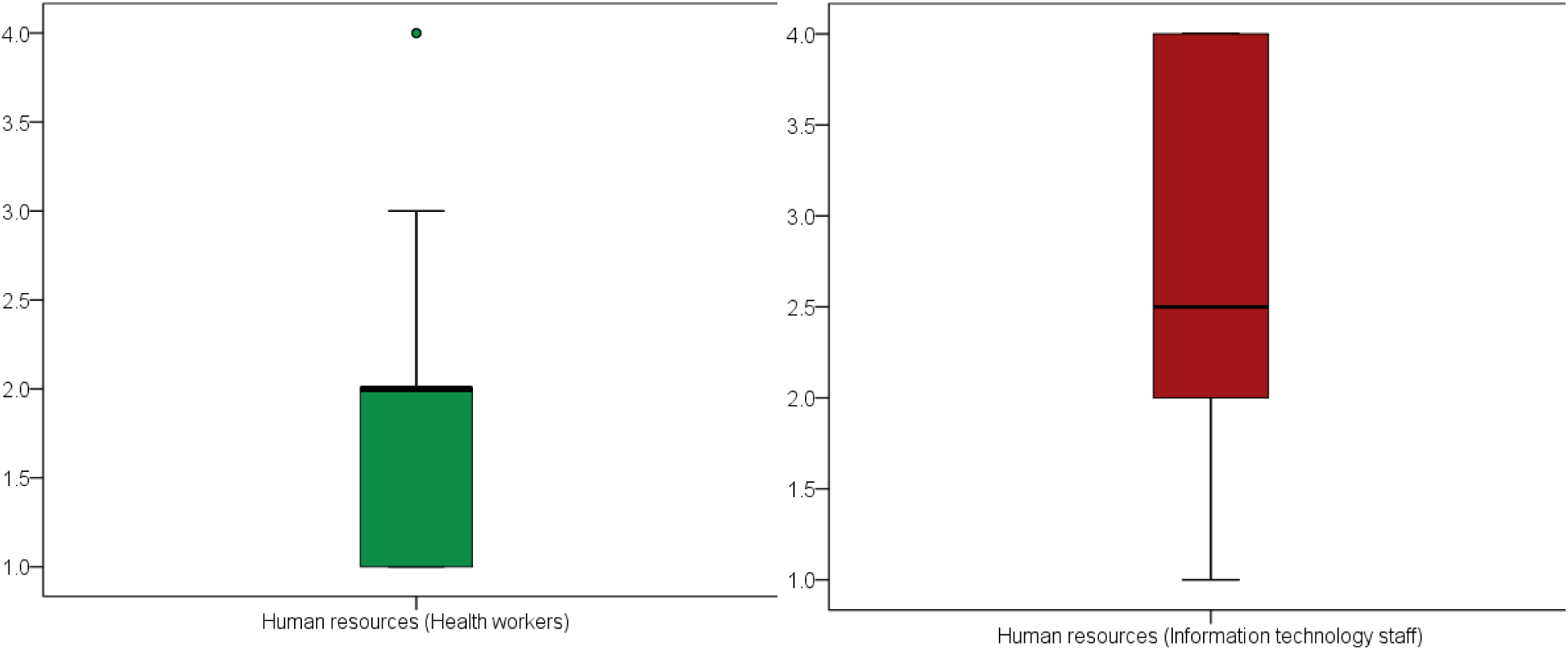
Maturity level of human resources (health workers and information technology staff) for telemedicine delivery. **Footnote:** Boxplots illustrating the median maturity level and interquartile range. Human resources boxplot includes 3 outlier institutions (green dot) with median of 4.0. Health workers (green boxplot) (Median 2.0 (1.0), range (1 – 4), mean (95% CI) (1.9 (1.5 – 2.4)); Information technology staff (wine boxplot) (Median 2.5 (2.0), range (1 – 4), mean (95% CI) (2.7 (2.3 – 3.2))

**Table 3.** Overall maturity of digital environment for delivery of telemedicine services

| Digital environment | Maturity level | Maturity level |
| --- | --- | --- |
|  | (Median (IQR)) | (Range) |
|  | n = 24 | n=24 |
| Internet connection and connectivity | 2.0 (1.9) | 1 – 4 |
| Software (managing medical records, patient portals) | 1.5 (1.0) | 1 – 3 |
| Administrative software (billing, payments, monitoring) | 2.0 (1.8) | 1 – 4 |
| Technical equipment (e.g., hardware, others) | 2.25 (1.0) | 1 – 4 |

## DISCUSSION

This is the first nationally representative exploratory survey documenting the maturity and level of preparedness of federally funded tertiary health institutions in Nigeria to deploy and integrate telemedicine services in healthcare provision. The objectives of our survey align with the World Health Organization’s recommendations that evaluation is vital for scalability, transferability, and continuing quality improvement of telemedicine.^7^ The main findings indicate a widely varied organizational readiness, predominantly beginner level with respect to the processes required for successful deployment of telemedicine services, the digital environment, human resources and expertise, and the lowest level of preparedness with respect to regulations.

### Organizational readiness

This examines the foundational issues that need to be resolved before an institution goes ahead to implement telemedicine services. In this study, the median maturity level for organizational readiness indicates that participating institutions are primarily at the beginner level to deploy telemedicine services, although considerably varied. The PAHO tool for assessing institutional maturity recommends delaying the implementation of telemedicine services if the institutional score is less than a maturity level of 3 for related items. Each institution in this category will need to formulate an action plan geared towards taking their institutuional readiness to a higher level in the areas identified as lacking (such as availability of IT staff, reliable internet, infrastructure for providing telemedicine services, etc.), before proceeding with telemedicine service provision. The main reason would be to avoid the frustration and failure that could result from launching telemedicine without the adequate capacity. In this study less than 20% of institutions had a budget line for telemedicine services, exposing a potential weakness that should be addressed as an area of priority. Not surprisingly, given the relative lack of funding, ∼ 1/3^rd^ of participating institutions had commenced any telemedicine services at the time of the survey. Improving institutional readiness would understandably involve a policy shift and the appropriate political will to divert financial resources to build capacity. In line with the general principles of quality improvement, it is recommended that facilities roll out services in phases, recognizing areas of critical or urgent need, where the impact would be most felt. ^8, 9^ This would also help to build confidence of both providers and users in telemedicine and increase buy-in to the concept of e-health, allowing for a period of transition in deploying and up-scaling the technology and user capabilities required for telemedicine service provision.^8, 10^

### Processes

The median maturity score reflecting the set of operations and functions required to commence telemedicine services implied there were no initiatives in place. Key considerations in this area include assignment of duties to clinical and administrative staff, security, procedures for referrals, adverse events, and emergency support.

Without the recommended processes in place, telemedicine services are not likely to be sustainable, or would quickly encounter clinical, technical or legal operational challenges capable of eroding confidence and user acceptability. We recommend that institutions include training and reasonable timelines for developing and rolling out actions related to relevant processes, and tasking specified working groups with sufficient expertise to midwife the processes. Staff training has been identified as one of the prerequisite measures for sustainability of any successful e-health system, and the fundamental role of collaboration for capacity building to ensure success and sustainability is a key lesson reiterated by the findings from previous global surveys. ^7, 10^

### Digital environment

Digitalization of processes and robust information and communications technology (ICT) infrastructure are fundamental requirements for efficient telemedicine services, although these should be relatively adaptable to the level of technological advancement, cognizant of appropriateness of solutions to local context, optimize cost-effectiveness, and minimize complexity. ^4, 7, 10^ The digital environment (technology infrastructure) to support deployment of telemedicine was assessed in this study to be at a median beginner level across participating institutions, ranging from less than beginner level for patient records software (median 1.5) to slightly above beginner level for technical equipment. This observation is practically actionable and can be addressed with budgetary allocations and manpower development specifically targeted at installing robust health information management systems (incorporating administrative applications, clinical software and hardware) integrating a platform for telemedicine.

Although Nigeria is considered to have increased its telecommunications coverage in the last several years,^11^ other related infrastructure such as challenges of interruptions in power supply, limited internet bandwidth and data costs are cogent barriers to implementation of telemedicine on a larger scale, including migration to real-time synchronous access rather than store-and –forward asynchronous interfaces. ^9, 10, 12, 13^ This challenge is similar across less developed countries and impacts providers and users, as active mobile broadband subscriptions, proportion of individuals using the internet, and mobile cellular telephone subscriptions are approximately 33%, 19% and 75% (compared to corresponding rates of 122%, 87% and 129% respectively in high income countries).^14^ According to the 2020 report of the International Telecommunications Unit (ITU), about a quarter of less developed countries lack access to a mobile-broadband network (more so in the rural than urban settings), coming short of the sustainable development goal (SDG) (9.c) to significantly increase access to ICT and strive to provide universal and affordable access to the Internet in least developed countries by 2020.^14^ For telemedicine services to achieve full potential in Nigeria, the overall national status of ICT development has to improve.

### Human Resources

The human resources requirement for implementing and sustaining telehealth services include IT, administrative and health workforce, and long-term commitment to developing this component is an important investment for achieving sustainable telemedicine services. ^7, 10^ Committing resources to and implementing well-coordinated e-health training derived from evidence-based conceptual frameworks recognizes the importance of human capacity in telehealth and also avoids the frustration of unstructured and random strategies at capacity building with a high likelihood of being unsustainable. ^15, 16^ In our survey, most institutions self-reported being at a beginner level in this regard although all reported having taken some steps to provide qualified IT personnel for telemedicine. It is entirely conceivable that without any concerted and proactive intervention, the current out-migration of the specialized health workforce and IT personnel being experienced as part of the current wave of ‘brain-drain’ potentially will further jeopardize institutional capacity to advance in this domain.

### Regulation

The institutional maturity vis-à-vis regulatory requirements for telemedicine was the least mature, with majority indicating being least ready. This observation was a common thread even in other segments of the survey where a few questions bordering on legal or regulatory issues were encountered. For example, less than 20% of institutions were at advanced levels of maturity with regard to knowledge/clarity of all the legal issues pertaining to telemedicine, as well as issues relating to what constitutes malpractice.

Other studies report a lack of legal framework to cover telemedicine service delivery in their facility/locality within Nigeria,^12^ and a number of studies report staff concerns about medico-legal implications of telemedicine, issues of data privacy, impersonation and quackery.^17^ A well-structured regulatory framework is crucial for a successful and sustainable telemedicine service. The 2010 WHO report on the second global survey on eHealth noted that lower-middle and low-income countries (LMICs) had the lowest percentage of countries with policies or legislation to define medical jurisdiction, liability, reimbursement, patient safety and quality of care, and protection of privacy of personally identifiable information and privacy of individual’s electronic health data.

Moreover, policy related considerations constituted the third highest barrier to telemedicine in LMICs (compared to being the seventh highest for high income counties). ^7^

### Expertise

Under this domain, the median level of maturity was 2.0 (IQR=2.0). This category summarizes the political will required to sustain telemedicine after implementation. It is the drive that the institutions require for the telemedicine services to run successfully, and incorporates tools that pertain more to the overall effect of telemedicine services on public health. A global survey on telemedicine utilization for movement disorders during the COVID-19 pandemic observed an increase in the utilization of different forms of telemedicine, including in Nigeria, in a bid to reduce spread of the virus whilst still providing some degree of care to patients; a scenario that cut across almost all fields of medicine. ^12, 18, 19^ Prior to this, in 2010, approximately 25% of LMICs cited expertise as a barrier to telemedicine (in contrast to about 5% of HICs).^7^ This demonstrates that expertise is rapidly up-scalable where an urgent need is recognized, and barriers cited as anchors for not using telemedicine are surmountable.

Any telemedicine service design should not just focus on the “technology” but should employ a holistic approach that will, in addition to the technological capabilities, encompass changes in clinical processes, clinical governance, change management, clinician and patient expectations, and operational sustainability.^20^ A recent systematic review on telemedicine use in sub-Saharan Africa concluded that while policy remains the same across the region, policy makers need to develop implementation strategies in tune with the needs of the individual countries.^21^ The authors recommended establishment of relevant policies, legislations and ethical standards to govern telemedicine use in healthcare in the region.^21^ This is particularly relevant for Nigeria as there is currently no regulatory agency for telemedicine-related matters.^22, 23^ From the legal perspective, there has been a call for enactment of legislation for telemedicine in Nigeria and a need for advocacy for a legal framework for telemedicine.^23^

In conclusion, our survey demonstrates the current status of telemedicine maturity of our federally funded tertiary health institutions that are at the fore of healthcare delivery and manpower training in Nigeria. We recommend that policy makers re-evaluate the commitment to implementing the tripartite strategic objectives of the Global Strategy on Digital Health (2020-2025) including promoting global collaboration and advancing the transfer of knowledge on digital health, advancing the implementation of national digital health strategies, and strengthening governance of digital health at the national level.^24^ These strategies promote the pillars of sustainable telehealth delivery: infrastructure and appropriate technology, policy and governance, manpower and capacity development.^7, 24^ Such a comprehensive approach will assure sustainability of any existing and future telemedicine programmes in the country.

### Limitations

We acknowledge the main limitation of the study regarding the self-report nature of the data provided as we did not conduct an actual facility inspection to verify the responses. However, we sourced responses from the administrative leadership at each institution to improve the veracity and credibility of the information. Our data are largely representative of the scenario across the nation, with the understanding that specific institutional status would vary within the sample studied, and as expected, at specific institutions not included in the survey. Notwithstanding, the geographic spread and the similarity of the profile of institutions included to other federally funded tertiary institutions across the country justifies the relevance of the findings and recommendations emanating thereof.

## Data Availability

All data produced in the present work are contained in the manuscript

---

**Supplementary Table 1.** PAHO Telemedicine maturity levels of federally funded tertiary health institutions in Nigeria (Organizational readiness)

| Parameters/questions from PAHO toolkit | Median maturity level (IQR) | Levels of maturity n (%) |  |  |  | n requiring technical support (%) |
| --- | --- | --- | --- | --- | --- | --- |
| I. ORGANIZATIONAL READINESS |  | 1 | 2 | 3 | 4 |  |
| Is senior management committed to offering telemedicine services? | 3.0 (1.0) | 1 (4.3) | 7 (30.4) | 8 (34.8) | 7 (30.40) | 2 (8.3) |
| Is it clearly understood which services can be offered via telemedicine? * | 2.0 (1.0) | 4 (17.4) | 6 (26.1) | 7 (30.4) | 6 (26.1) | 3 (12.5) |
| Have the services to be offered via telemedicine been identified? * | 2.0 (2.0) | 6 (27.3) | 6 (27.3) | 8 (36.4) | 2 (9.1) | 5 (20.8) |
| Is there a budget available for offering telemedicine services? * | 1.0 (0.0) | 18 (78.3) | 4 (17.4) | 0 (0) | 1 (4.3) | 4 (16.7) |
| Is the IT staff trained to provide support services for telemedicine?* | 2.2 (2.0) | 5 (22.7) | 8 (36.4) | 3 (13.6) | 6 (27.3) | 7 (29.2) |
| Do the national or local regulations allow the implementation of telemedicine services? | 3.0 (2.0) | 8 (33.3) | 3 (12.5) | 6 (25.0) | 7 (29.2) | 2 (8.3) |
| Does the institution have reliable internet access? * | 2.5 (2.0) | 4 (18.2) | 5 (22.7) | 8 (36.4) | 5 (22.7) | 5 (20.8) |
| Does the institution already have some kind of telemedicine program in operation? | 2.0 (2.0) | 9 (37.5) | 7 (29.2) | 2 (8.3) | 6 (25.0) | 5 (20.8) |
| Does the institution have any experience in using instant messaging or texting for health promotion? | 2.0 (2.0) | 9 (37.5) | 6 (25.0) | 5 (20.8) | 4 (16.7) | 5 (20.8) |
| Does the institution have any experience in health service delivery through virtual consultations? * | 1.5 (1.0) | 9 (39.1) | 6 (26.1) | 1 (4.3) | 7 (30.4) | 6 (25.0) |
| Does the institution have any experience in remote monitoring of patients? * | 1.0 (1) | 11 (47.8) | 7 (30.4) | 3 (17.4) | 1 (4.3) | 4 (16.7) |
| Parameters/questions from PAHO toolkit | Median | Levels of maturity |  |  |  | n |

|  | maturity<br>level (IQR) | n (%)<br>n requiring technical support (%) |  |  |  | requiring<br>technical<br>support<br>(%) |
| --- | --- | --- | --- | --- | --- | --- |
| **I. ORGANIZATIONAL READINESS** |  | **1** | **2** | **3** | **4** |  |
| Could the funding of telemedicine services be extended beyond the initial planning and pilot stages, to become a sustainable model? * | 3.0 (2) | 6 (26.1) | 2 (8.7) | 7 (30.4) | 8 (34.8) | 3 (12.5) |
| Is medical staff trained on providing telemedicine services? See section IV (on human resources) below for more information. * | 1.0 (1) | 13 (56.5) | 8 (33.3) | 1 (4.3) | 1 (4.3) | 4 (16.7) |
| If the score given on the previous question is 1 or 2, have telemedicine training options been determined? | 1.0 (2) | 14 (66.7) | 2 (9.5) | 4 (19.0) | 1 (4.8) | 3 (12.5) |
| Does the institution have the infrastructure necessary for providing telemedicine services? | 2.5 (2) | 3 (12.5) | 7 (29.2) | 5 (20.8) | 9 (37.5) | 5 (20.8) |
| • Sufficient space | 2.5 (1) | 3 (12.5) | 6 (25.0) | 8 (33.3) | 7 (29.2) | 0 (0.0) |
| • Reliable electricity supply | 3.0 (2) | 2 (8.3) | 4 (16.7) | 5 (20.8) | 13 (54.2) | 0 (0.0) |
| • Acceptable illumination levels | 3.0 (2) | 2 (8.3) | 6 (25.0) | 7 (29.2) | 9 (37.5) | 1 (4.2) |
| • Support teams | 3.0 (1) | 3 (13.0) | 6 (26.1) | 9 (39.1) | 5 (21.7) | 5 (20.8) |
| Has anyone been designated to be responsible for telemedicine services? | 1.0 (1) | 13 (54.2) | 6 (25.0) | 3 (12.5) | 2 (8.3) | 3 (12.5) |
| Is medical staff in agreement with offering telemedicine services? | 2.0 (2) | 7 (29.2) | 5 (20.8) | 8 (33.3) | 4 (16.7) | 1 (4.2) |
| Has anything been done to address resistance to changes in routines with which physicians feel safe and comfortable, and to a new and unknown system involving a certain degree of initial uncertainty? | 1.5 (1) | 10 (41.7) | 10 (41.7) | 2 (8.3) | 2 (8.3) | 1 (4.2) |
| If the score given on the previous question is 1 or 2, can this problem be solved through participatory dialogue? * | 3.0 (2) | 1 (5.3) | 4 (21.1) | 8 (42.1) | 6 (31.6) | 1 (4.2) |
| Have any incentives been put in place to promote the use of telemedicine? * | 1.0 (0) | 21 (95.5) | 1 (4.5) | 0 (0.0) | 0 (0.0) | 3 (12.5) |
| Is medical staff familiar with privacy and security practices based on current ethical and legal principles? | 2.0 (1) | 5 (20.8) | 11 (45.8) | 5 (20.8) | 3 (12.5) | 3 (12.5) |
| Has the workload involved in launching this type of program in the current environment been defined? | 1.0 (1) | 13 (54.2) | 6 (25.0) | 4 (16.7) | 1 (4.2) | 2 (8.3) |
| Does the program have the support of an institution specialized in telemedicine services? * | 1.5 (2) | 11 (47.8) | 6 (26.1) | 4 (17.4) | 2 (8.7) | 1 (4.2) |
| Has the institution's staff been informed of the intention to implement or expand telemedicine services? | 1.5 (2) | 9 (37.5) | 5 (20.8) | 6 (25.0) | 4 (16.7) | 1 (4.2) |

| Parameters/questions from PAHO toolkit | Median maturity level (IQR) | Levels of maturity<br>n (%)<br>n requiring technical support (%) |  |  |  | n requiring technical support (%) |
| --- | --- | --- | --- | --- | --- | --- |
|  |  | 1 | 2 | 3 | 4 |  |
| **I. ORGANIZATIONAL READINESS** |  |  |  |  |  |  |
| Have the potential beneficiaries of these telemedicine services been informed of their launch or expansion? | 1.0 (1) | 13 (54.2) | 7 (29.2) | 2 (8.3) | 2 (8.3) | 1 (4.2) |
| Have patient care agendas been changed due to the need for remote consultations? | 1.0 (0) | 18 (75.0) | 4 (16.7) | 1 (4.2) | 1 (4.2) | 2 (8.3) |
| What is the expected level of acceptance for telemedicine services from their potential beneficiaries? | 2.0 (3) | 5 (20.8) | 7 (29.2) | 5 (20.8) | 7 (29.2) | 1 (4.2) |
| Are there cultural or linguistic barriers that could cause difficulties during the delivery of telemedicine services? | 1.5 (2) | 9 (37.5) | 7 (29.2) | 3 (12.5) | 5 (20.8) | 1 (4.2) |
| Is the level of connectivity of potential patients known? | 1.0 (0) | 17 (70.8) | 6 (25.0) | 0 (0) | 1 (4.2) | 4 (16.7) |
| Is the level of digital literacy of potential patients known? | 1.0 (0) | 15 (62.5) | 6 (25.0) | 2 (8.3) | 1 (4.2) | 4 (16.7) |
| Have governance mechanisms been established? | 1.0 (0) | 17 (73.9) | 3 (12.5) | 2 (8.7) | 1 (4.3) | 5 (2.8) |
| Have continuous assessment mechanisms been established? * | 1.0 (0) | 17 (73.9) | 4 (17.4) | 2 (8.7) | 0 (0) | 5 (20.8) |
| Are there formal procedures to remotely obtain informed consent from patients? * |  |  |  |  |  |  |

**Footnote: \* shows number of item non-responses. Available case analysis used.**

**Supplementary Table 2.** PAHO Telemedicine maturity levels of federally funded tertiary health institutions in Nigeria (Processes)

| Parameters/questions from PAHO toolkit | Median maturity level (IQR) | Levels of maturity n (%) |  |  |  | n requiring technical support (%) |
| --- | --- | --- | --- | --- | --- | --- |
| II. PROCESSES |  | 1 | 2 | 3 | 4 |  |
| Have the duties, roles, and responsibilities associated with the telemedicine services been defined for all staff who will be involved?* | 1.0 (1.0) | 13 (56.5) | 8 (34.8) | 1 (4.3) | 1 (4.3) | 6 (25.0) |
| Have the duties, roles, and responsibilities related to the telemedicine services been defined for all administrative staff? * | 1.0 (1.0) | 15 (65.2) | 5 (21.7) | 2 (8.7) | 1 (4.3) | 6 (25.0) |
| Have procedures been defined to address the security considerations regarding patients and institutional legal responsibility? * | 1.0 (0.0) | 18 (78.3) | 3 (13.0) | 1 (4.3) | 1 (4.3) | 6 (25.0) |
| Have procedures been defined to safeguard the security, confidentiality, and backup of the data and information generated by telemedicine services? | 1.0 (1.0) | 17 (70.8) | 4 (16.7) | 1 (4.2) | 2 (8.3) | 5 (20.8) |
| Are procedures in place to record the level of patient satisfaction with telemedicine services? * | 1.0 (1.0) | 16 (66.7) | 5 (20.8) | 2 (8.3) | 1 (4.2) | 6 (25.0) |
| Are notification procedures in place in the event of incidents or adverse events during telemedicine consultations? * | 1.0 (0.0) | 18 (78.3) | 2 (8.7) | 3 (13.0) | 0 (0.0) | 6 (25.0) |
| Are there standardized procedures to notify and document possible technical failures during a consultation that could affect the clinical outcome? * | 1.0 (0.0) | 18 (78.3) | 5 (21.7) | 0 (0.0) | 0 (0.0) | 6 (25.0) |
| Are there formal procedures to remotely obtain informed consent from patients? * | 1.0 (0.0) | 19 (79.2) | 4 (16.7) | 0 (0.0) | 0 (0.0) | 6 (25.0) |
| Are there procedures or tools for staff and patients to share their concerns, suggestions, or comments regarding how the telemedicine program is working? ** | 1.0 (0.0) | 18 (81.8) | 3 (13.6) | 1 (4.5) | 0 (0.0) | 4 (16.7) |
| Is there a strategy and operation plan to orient healthcare providers to-wards opting for outpatient teleconsultations and remote monitoring of their patients? | 1.0 (1.0) | 17 (70.8) | 2 (8.3) | 4 (16.7) | 1 (4.2) | 4 (16.7) |
| Are there communication mechanisms for informing and educating | 1.0 (1.0) | 16 (66.7) | 4 (16.7) | 1 (4.2) | 3 (12.5) | 4 (16.7) |
| the public about the recommended use of telemedicine? |  |  |  |  |  |  |
| Is there a procedure or emergency plan for physicians practicing telemedicine if they consider that a patient should be referred to an intensive care center? | 1.0 (1.0) | 15 (62.5) | 4 (16.7) | 2 (8.3) | 3 (12.5) | 4 (16.7) |

**Footnote: \* shows number of item non-responses. Available case analysis used.**

**Supplementary Table 3.** PAHO Telemedicine maturity levels of federally funded tertiary health institutions in Nigeria (Digital environment)

| Parameters/questions from PAHO toolkit | Median maturity level (IQR) | Levels of maturity n (%) |  |  |  | n requiring technical support (%) |
| --- | --- | --- | --- | --- | --- | --- |
| III.DIGITAL ENVIRONMENT |  |  |  |  |  |  |
| Internet connection and connectivity |  | 1 | 2 | 3 | 4 |  |
| Is a regular, reliable internet connection available? * | 3 (2) | 3 (13.0) | 7 (30.4) | 7 (30.4) | 6 (25.0) | 5 (20.8) |
| Does the available bandwidth make it possible to offer telemedicine services without affecting other services? * | 2.5 (1) | 5 (21.7) | 8 (34.8) | 7 (30.4) | 3 (13.0) | 5 (20.8) |
| Is the institution able to calculate the bandwidth necessary for providing telemedicine services?* | 2 (2) | 11 (47.8) | 6 (26.1) | 4 (17.4) | 2 (8.7) | 6 (25.0) |
| Is the necessary minimum hardware available?* | 3 (2) | 4 (17.4) | 9 (39.1) | 5 (21.7) | 5 (26.3) | 5 (20.8) |
| If the score given on the previous question is 1 or 2, is there a budget available to procure the necessary equipment? | 1 (2) | 6 (25.0) | 7 (29.2) | 1 (4.2) | 0 (0) | 3 (12.5) |
| Is there technical support available to solve problems related to connectivity? ** | 2 (2) | 2 (8.7) | 10 (43.4) | 4 (17.4) | 6 (26.1) | 4 (16.7) |
| Is there a cybersecurity plan?* | 1 (2) | 13 (56.5) | 4 (17.4) | 6 (26.1) | 0 (0) | 5 (20.8) |
| Are technical support manuals available to address connectivity issues? *** | 2 (2) | 10 (47.6) | 6 (28.6) | 3 (14.3) | 2 (9.5) | 7 (29.2) |
| Is there a contingency plan in the event of equipment or connectivity failures? *** | 1.5 (1) | 9 (42.9) | 8 (38.1) | 2 (9.5) | 2 (9.5) | 7 (29.2) |
| Has the possible impact of the new telemedicine services on the current technological infrastructure been considered?* | 1 (1) | 13 (56.5) | 6 (26.1) | 2 (8.7) | 2 (8.7) | 2 (8.3) |

| Applications (software for managing medical records, patient portals, etc) | Median maturity level (IQR) | Levels of maturity n (%) |  |  |  | n requiring technical support (%) |
| --- | --- | --- | --- | --- | --- | --- |
|  |  | 1 | 2 | 3 | 4 |  |
| Is there an electronic patient record system? | 3 (2) | 2 (8.3) | 8 (33.3) | 8 (33.3) | 6 (25.0) | 2 (8.3) |
| Is there a patient portal? | 1.5 (2) | 12 (50.0) | 6 (25.0) | 2 (8.3) | 4 (16.7) | 3 (12.5) |
| Does the institution know what software or IT solutions are necessary to offer telemedicine services?*** | 2 (1) | 8 (38.1) | 8 (38.1) | 2 (9.5) | 3 (14.3) | 6 (25.0) |
| Are there standard operating procedures for managing the data and processes involved in telemedicine delivery?* | 1 (1) | 14 (60.9) | 6 (26.1) | 2 (8.7) | 1 (4.3) | 5 (20.8) |
| Are there guidelines on patient safety?* | 1.5 (2) | 11 (47.8) | 5 (21.7) | 6 (26.1) | 1 (4.3) | 3 (12.5) |
| Are there guidelines on privacy and data confidentiality?* | 1.5 (1) | 11 (47.8) | 7 (30.4) | 4 (17.4) | 1 (4.3) | 3 (12.5) |
| What is the level of interoperability among the telemedicine services' different systems and databases?* | 1 (1) | 16 (69.6) | 6 (26.1) | 1 (4.3) | 0 (0) | 4 (16.7) |
| Are there terms of reference for the procurement of IT solutions?* | 2 (2) | 10 (43.5) | 7 (30.4) | 3 (13.0) | 3 (13.0) | 3 (12.5) |
| Has it been decided whether telemedicine IT solutions will be integrated into other existing systems and processes, such as those for medical records, patient portals, and messaging? | 1 (1) | 13 (54.2) | 6 (25.0) | 3 (12.5) | 2 (8.3) | 4 (16.7) |
| Are there standard operating procedures for managing data and processes related to providing care to patients? | 2 (2) | 8 (33.3) | 9 (37.5) | 5 (20.8) | 2 (8.3) | 3 (12.5) |
| Do the medical records platforms used in the telemedicine services have the capacity to include copies of all the electronic communications related to the patient? ** | 1 (1) | 15 (68.2) | 4 (18.2) | 3 (13.6) | 0 (0) | 5 (20.8) |
| **Applications (administrative software for billing, payments, monitoring hours, etc.)** | **Median maturity level** | **Levels of maturity n (%)** |  |  |  | **n requiring** |

|  | (IQR | 1 | 2 | 3 | 4 | technical support (%) |
| --- | --- | --- | --- | --- | --- | --- |
| Are the administrative management platforms ready to work with the implementation of telemedicine services? | 2 (2) | 6 (25.0) | 10 (41.7) | 3 (12.5) | 5 (20.8) | 4 (16.7) |
| **Technical equipment (hardware & other equipment)** | **Median maturity level (IQR** | **1** | **2** | **3** | **4** | **n requiring technical support (%)** |
| Is there an inventory of all the technical equipment, including the brand, model, time in operation, and serial number? * | 2 (1) | 5 (21.7) | 13 (56.5) | 3 (13.0) | 2 (8.7) | 4 (16.7) |
| Is there a secure location to store the equipment when is it not in use? | 3 (1) | 2 (8.3) | 7 (29.2) | 10 (41.7) | 5 (20.8) | 1 (4.2) |
| Is there a maintenance program for the technical equipment? * | 2 (2) | 7 (30.4) | 7 (30.4) | 7 (30.4) | 2 (8.7) | 5 (20.8) |
| Has consideration been given to the technological capacity for storage and security that will be necessary to document and record face-to-face meetings? * | 2 (1) | 7 (30.4) | 12 (52.2) | 2 (8.7) | 2 (8.7) | 6 (25.0) |
| Is technical support available from IT specialists? * | 3 (1) | 3 (13.0) | 5 (21.7) | 11 (47.8) | 4 (17.4) | 5 (20.8) |
| Are there plans for updating technical equipment? | 3 (1) | 4 (16.7) | 7 (29.2) | 8 (33.3) | 5 (20.8) | 3 (12.5) |

**Footnote: \* shows number of item non-responses. Available case analysis used.**

**Supplementary Table 4.**
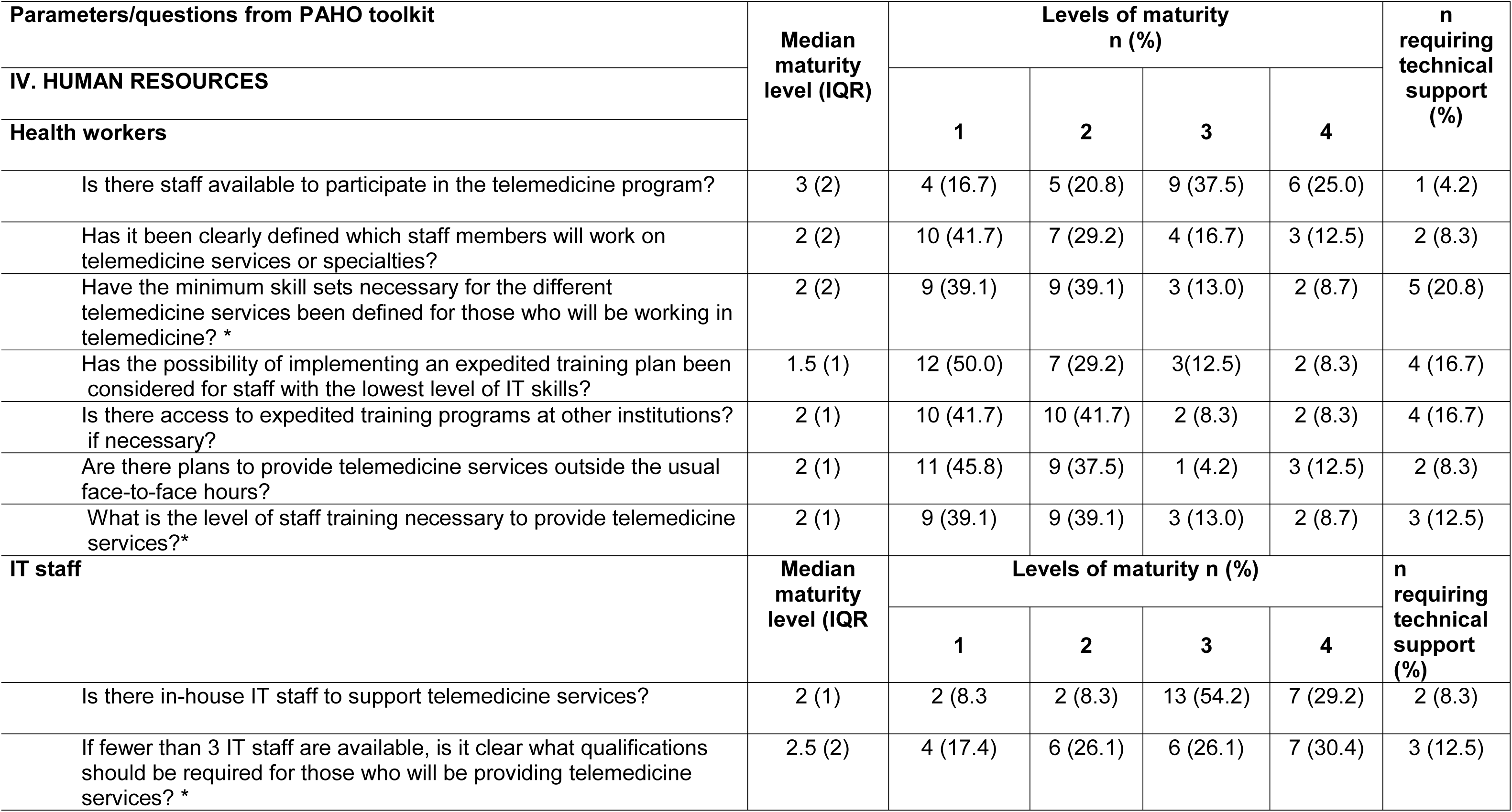
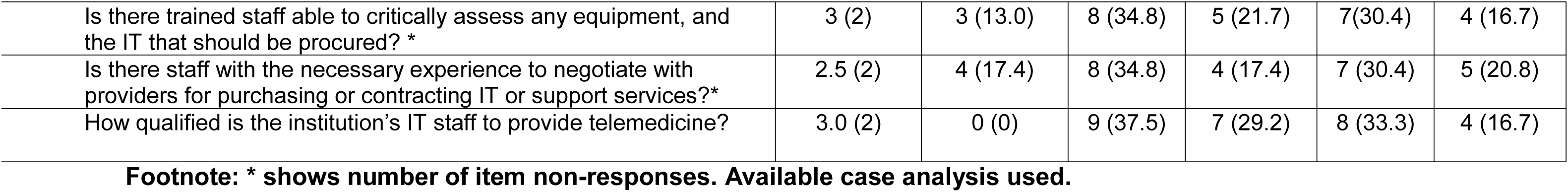
PAHO Telemedicine maturity levels of human resources of federally funded tertiary health institutions in Nigeria (Human resources)

| Parameters/questions from PAHO toolkit | Median maturity level (IQR) | Levels of maturity n (%) |  |  |  | n requiring technical support (%) |
| --- | --- | --- | --- | --- | --- | --- |
| IV. HUMAN RESOURCES |  | 1 | 2 | 3 | 4 |  |
| **Health workers** |  |  |  |  |  |  |
| Is there staff available to participate in the telemedicine program? | 3 (2) | 4 (16.7) | 5 (20.8) | 9 (37.5) | 6 (25.0) | 1 (4.2) |
| Has it been clearly defined which staff members will work on telemedicine services or specialties? | 2 (2) | 10 (41.7) | 7 (29.2) | 4 (16.7) | 3 (12.5) | 2 (8.3) |
| Have the minimum skill sets necessary for the different telemedicine services been defined for those who will be working in telemedicine? * | 2 (2) | 9 (39.1) | 9 (39.1) | 3 (13.0) | 2 (8.7) | 5 (20.8) |
| Has the possibility of implementing an expedited training plan been considered for staff with the lowest level of IT skills? | 1.5 (1) | 12 (50.0) | 7 (29.2) | 3 (12.5) | 2 (8.3) | 4 (16.7) |
| Is there access to expedited training programs at other institutions? if necessary? | 2 (1) | 10 (41.7) | 10 (41.7) | 2 (8.3) | 2 (8.3) | 4 (16.7) |
| Are there plans to provide telemedicine services outside the usual face-to-face hours? | 2 (1) | 11 (45.8) | 9 (37.5) | 1 (4.2) | 3 (12.5) | 2 (8.3) |
| What is the level of staff training necessary to provide telemedicine services?* | 2 (1) | 9 (39.1) | 9 (39.1) | 3 (13.0) | 2 (8.7) | 3 (12.5) |
| **IT staff** | Median maturity level (IQR) | Levels of maturity n (%) |  |  |  | n requiring technical support (%) |
|  |  | 1 | 2 | 3 | 4 |  |
| Is there in-house IT staff to support telemedicine services? | 2 (1) | 2 (8.3) | 2 (8.3) | 13 (54.2) | 7 (29.2) | 2 (8.3) |
| If fewer than 3 IT staff are available, is it clear what qualifications should be required for those who will be providing telemedicine services? * | 2.5 (2) | 4 (17.4) | 6 (26.1) | 6 (26.1) | 7 (30.4) | 3 (12.5) |
| Is there trained staff able to critically assess any equipment, and the IT that should be procured? * | 3 (2) | 3 (13.0) | 8 (34.8) | 5 (21.7) | 7(30.4) | 4 (16.7) |
| Is there staff with the necessary experience to negotiate with providers for purchasing or contracting IT or support services?* | 2.5 (2) | 4 (17.4) | 8 (34.8) | 4 (17.4) | 7 (30.4) | 5 (20.8) |
| How qualified is the institution's IT staff to provide telemedicine? | 3.0 (2) | 0 (0) | 9 (37.5) | 7 (29.2) | 8 (33.3) | 4 (16.7) |

**Footnote: \* shows number of item non-responses. Available case analysis used.**

**Supplementary Table 5.** PAHO Telemedicine maturity levels of federally funded tertiary health institutions in Nigeria (Regulatory Issues)

| Parameters/questions from PAHO toolkit | Median maturity level (IQR) | Levels of maturity n (%) |  |  |  | n requiring technical support (%) |
| --- | --- | --- | --- | --- | --- | --- |
| **V. REGULATORY ISSUES** |  | **1** | **2** | **3** | **4** |  |
| Are all the legal issues associated with the delivery of telemedicine services perfectly clear? ** | 1.0 (1.0) | 15 (65.2) | 6 (26.1) | 2 (8.7) | 0 (0.0) | 4 (16.7) |
| Is there a procedure in place to keep staff who provide telemedicine services up to date on possible changes in regulations, statutes, federal and subnational policies, and legislation related to telemedicine services? | 1.0 (1.0) | 13 (54.2) | 7 (29.2) | 4 (16.7) | 0 (0.0) | 4 (16.7) |
| Does the institution have in-house legal advisory services? Does it have access to a specialized consultancy to receive expert advice on legal, ethical, privacy, and security issues? | 3.0 (2.0) | 5 (20.8) | 6 (25.0) | 8 (33.3) | 5 (20.8) | 3 (12.5) |
| Is it known for certain that the patients are located within the same catchment area (e.g. state, province, or municipality) as the institution providing the telemedicine services? | 2.0 (2.0) | 10 (41.7) | 5 (20.8) | 6 (25.0) | 3 (12.5) | 3 (12.5) |
| Are the malpractice issues related to the telemedicine services well understood? | 1.0 (1.0) | 11 (45.8) | 9 (37.5) | 2 (8.3) | 2 (8.3) | 5 (20.8) |
| Has a process been put in place for obtaining and documenting the consent of patients before they participate in a telemedicine visit? * | 1.0 (1.0) | 14 (60.9) | 5 (21.7) | 3 (13.0) | 1 (4.3) | 6 (25.0) |
| Is there a regulatory framework for authorizing, integrating, and reimbursing telemedicine in the delivery of care to all patients, particularly during emergencies and outbreaks?* | 1.0 (0.0) | 17 (73.9) | 4 (17.4) | 2 (8.7) | 0 (0.0) | 6 (25.0) |
| Are new regulations or legal technical frameworks necessary to implement telemedicine services? ** | 2.0 (1.0) | 10 (45.5) | 7 (31.8) | 1 (4.5) | 4 (18.2) | 8 (33.3) |

**Footnote: \* shows number of item non-responses. Available case analysis used.**

**Supplementary Table 6.**
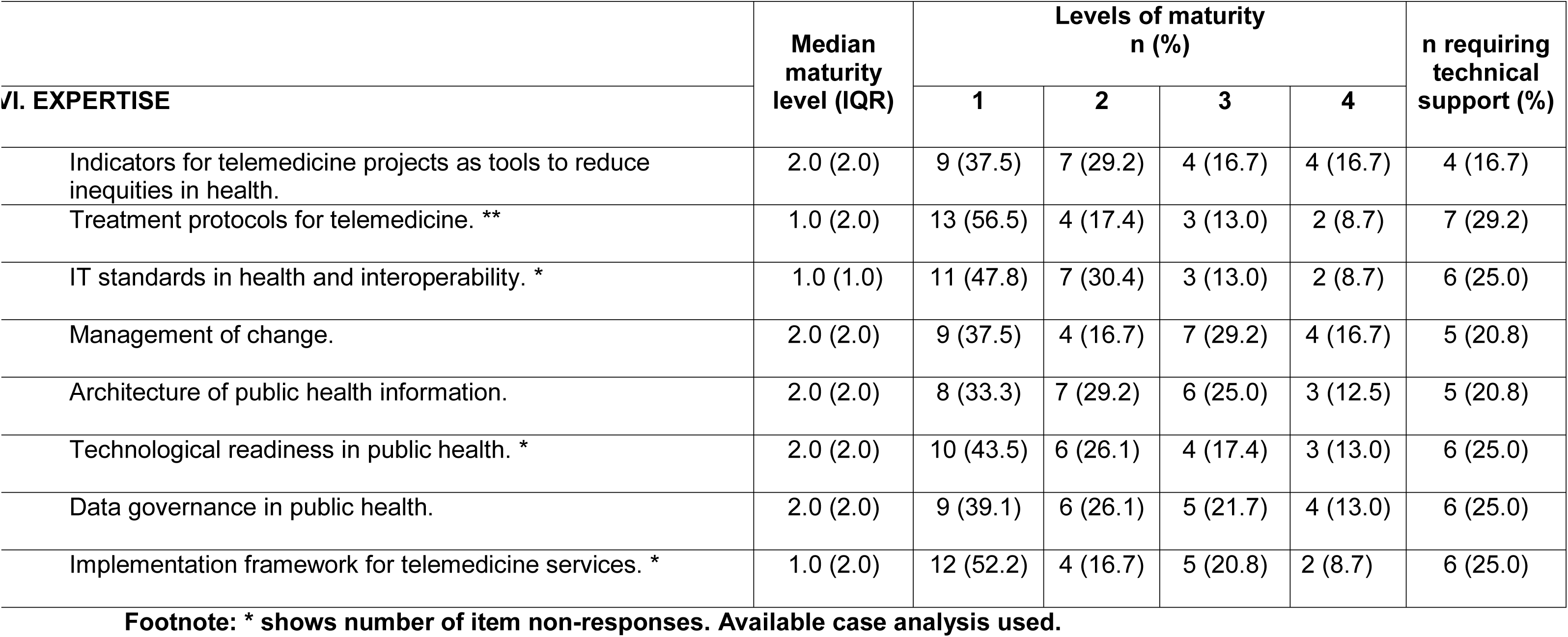
PAHO Telemedicine maturity levels of federally funded tertiary health institutions in Nigeria (Expertise)

|  | Median maturity level (IQR) | Levels of maturity n (%) |  |  |  | n requiring technical support (%) |
| --- | --- | --- | --- | --- | --- | --- |
| **VI. EXPERTISE** |  | **1** | **2** | **3** | **4** |  |
| Indicators for telemedicine projects as tools to reduce inequities in health. | 2.0 (2.0) | 9 (37.5) | 7 (29.2) | 4 (16.7) | 4 (16.7) | 4 (16.7) |
| Treatment protocols for telemedicine. ** | 1.0 (2.0) | 13 (56.5) | 4 (17.4) | 3 (13.0) | 2 (8.7) | 7 (29.2) |
| IT standards in health and interoperability. * | 1.0 (1.0) | 11 (47.8) | 7 (30.4) | 3 (13.0) | 2 (8.7) | 6 (25.0) |
| Management of change. | 2.0 (2.0) | 9 (37.5) | 4 (16.7) | 7 (29.2) | 4 (16.7) | 5 (20.8) |
| Architecture of public health information. | 2.0 (2.0) | 8 (33.3) | 7 (29.2) | 6 (25.0) | 3 (12.5) | 5 (20.8) |
| Technological readiness in public health. * | 2.0 (2.0) | 10 (43.5) | 6 (26.1) | 4 (17.4) | 3 (13.0) | 6 (25.0) |
| Data governance in public health. | 2.0 (2.0) | 9 (39.1) | 6 (26.1) | 5 (21.7) | 4 (13.0) | 6 (25.0) |
| Implementation framework for telemedicine services. * | 1.0 (2.0) | 12 (52.2) | 4 (16.7) | 5 (20.8) | 2 (8.7) | 6 (25.0) |

**Footnote: \* shows number of item non-responses. Available case analysis used.**

